# A randomized controlled safety and feasibility trial of floatation-REST in anxious and depressed individuals

**DOI:** 10.1101/2023.05.27.23290633

**Authors:** McKenna M. Garland, Raminta Wilson, Wesley K. Thompson, Murray B. Stein, Martin P. Paulus, Justin S. Feinstein, Sahib S. Khalsa

## Abstract

**Background:** Reduced Environmental Stimulation Therapy via floatation (floatation-REST) is a behavioral intervention designed to attenuate exteroceptive sensory input to the nervous system. Pilot studies in anxious and depressed individuals demonstrated that single sessions of floatation-REST are safe, well-tolerated, and associated with acute anxiolysis. However, there is not sufficient evidence of the feasibility of floatation-REST as a repeated intervention.

**Methods:** We randomized 75 individuals with anxiety and depression to six sessions of floatation-REST in different formats (pool-REST or pool-REST preferred) or an active comparator (chair-REST). Feasibility was assessed via adherence rate to the assigned intervention, tolerability via duration of REST utilization and overall study dropout rate, and safety via incidence of serious or non-serious adverse events.

**Results:** Six-session adherence was 85% for pool-REST, 89% for pool-REST preferred, and 74% for chair-REST. Dropout rates did not differ significantly between the treatment conditions.

Mean session durations were consistently above 50 minutes, and when allowed to choose the duration and frequency, participants opted to float for an average of 75 minutes. There were no serious adverse events associated with any intervention. Positive experiences were endorsed more commonly than negative ones and were also rated at higher levels of intensity.

**Conclusions:** Taken together, six sessions of floatation-REST appear feasible, well-tolerated, and safe in anxious and depressed individuals. Floatation-REST induces positively-valenced experiences with few negative effects. Larger randomized controlled trials evaluating markers of clinical efficacy are warranted.

**Clinical Trial Registration Identifier:** NCT03899090

## Introduction

Reduced Environmental Stimulation Therapy via floatation (i.e., floatation-REST) is a behavioral intervention designed to systematically attenuate exteroceptive sensory input to the nervous system. Dating back to the 1950’s, REST was first introduced to assess how humans would react to low-level or monotonous stimulation of the nervous system, with subsequent investigations exploring REST as a therapeutic intervention for habit modification during smoking cessation or stress reduction [1]. The earliest forms of REST involved strict confinement to a bed in a dimly lit cubicle for days at a time (i.e., ‘chamber-REST’) [2] or full-body submersion in a vertical water tank with the assistance of a breathing helmet [3, 4]. Since then, several technological developments have considerably expanded the availability of different REST formats. Currently, the most common form of floatation-REST involves floating effortlessly in a shallow pool of water heated to skin temperature and saturated with Epsom salt to increase buoyancy (consequently, it is sometimes also called ‘pool-REST’). The environment is specially engineered to be lightproof, soundproof, and humidity and temperature-controlled, so that input from visual, auditory, olfactory, gustatory, thermal, tactile, vestibular, and proprioceptive channels is minimized, as is movement and speech [5]. Chair-REST has also emerged as an alternative form of REST [6, 7]. In this environment, individuals recline in an ergonomically engineered chair designed to take pressure off the spinal cord. Typically, these chairs are placed in dimly lit and quiet rooms, similar but not identical to what would be experienced during pool-REST. Our previous work has highlighted that although the degree of sensory attenuation of chair-REST is not as substantial as traditional pool-REST, single sessions of chair-REST can produce effects on negative affect, anxiety, stress, relaxation, serenity, and refreshment of a similar nature in clinically anxious individuals [7]. Additionally, chair-REST induced a similar though weaker pattern of neural changes in resting state functional connectivity as pool-REST [6]. Such “zero-gravity chairs” have garnered public attention and are now available in many large department and furniture stores. The availability and utilization of pool- REST has grown in recent years, with hundreds of recreational “float centers” opening across the world during the past decade [8]. Despite the increase in public interest and consumption of various forms of REST, there has been limited research investigating the feasibility of the technique, especially in clinical populations.

Preliminary insights into the tolerability and safety of floatation-REST in anxious and depressed individuals are restricted to a few studies. A wait-list control trial reported that individuals scoring high on self-reported measures of generalized anxiety were largely adherent to 12 sessions of floatation-REST [9]. Another study found that individuals with stress-related pain and burnout depression were able to complete up to 33 sessions of floatation-REST [10]. However, neither of these studies assessed adverse events that could have arisen during the repeated sessions of floatation-REST, nor did they formally evaluate intervention feasibility. In prior studies, most anxious individuals who were assigned fixed durations of pool-REST or chair-REST for 45 to 90 minutes in length tolerated this amount [5, 7, 9, 11], but the impact of flexible assignment on preference has not been investigated. Finally, our initial safety studies examining the effects of single sessions of pool-REST or chair-REST in clinically anxious individuals revealed no serious adverse events, suggesting that the brief intervention exposure was safe and well-tolerated [5, 7]. Regarding safety, it is important to note that there have been several deaths reported with the use of floatation tanks at recreational float centers associated with concurrent drug (i.e., ketamine) or alcohol use [12], suggesting the need for inclusion of substance use screening. There have also been reports of auditory and visual hallucinations during floatation-REST [13]. However, these are generally described in a positive light, and are infrequent [1, 5]. Other negative events that have been reported include occasional skin itchiness or dry mouth, as well as discomfort resulting from accidental introduction of saltwater to the eyes or open cuts [5, 14]. Despite the common utilization of floatation-REST by the public, and an emerging clinical trials literature, no studies have systematically evaluated the safety profile of this intervention across multiple sessions.

This study examined the feasibility, tolerability, and safety of floatation-REST as a repeatable intervention in clinically anxious and depressed individuals using a randomized design with an active comparator (chair-REST). Our primary outcome was feasibility. We hypothesized that feasibility would be reflected by 6-session adherence rates of 80% or above, a level similar to standard behavioral interventions for anxiety and depression [15, 16]. As an additional feasibility assessment, we evaluated for potential differences in credibility and expectancy between the assigned REST interventions. We predicted that all forms of floatation-REST would be well-tolerated, with low dropout rates (<20%) over the course of the intervention and 6-month follow-up period. We also predicted that individuals would remain in their assigned REST environment for the majority of the allotted duration of time. Based on the prior literature, we predicted that the REST interventions would be safe and associated with minimal adverse reactions. Finally, as an exploratory aim, we examined the impact of flexible assignment on floatation-REST preference in terms of the chosen frequency and duration of float sessions, in order to provide information to help optimize the design of future trials.

## Methods

### Participant Recruitment

75 treatment-seeking adults with anxiety and depression were recruited through the Laureate Institute for Brain Research’s participant databases ([17] see Supplement for sample size determination). Inclusion criteria included high levels of current anxiety (as measured by an OASIS score ≥ 6; [18]) and anxiety sensitivity (as measured by an ASI-3 score ≥ 24; [19]). Of note, the initial eligibility criteria of OASIS score ≥ 8 and ASI-3 score ≥ 29 were lowered in September 2020 to increase participant recruitment to meet the targeted rate specified in the funded grant. Depression symptoms were measured via the Patient Health Questionnaire (PHQ- 9, [20]), but an inclusion cutoff score was not specified. Other inclusion criteria included: age between 18 and 60 years, and either no prior experience of floatation-REST or a minimum of 1 year since the last float session. If taking psychiatric medications or receiving psychotherapy, the treatment regimen was required to be stable, defined as having taken the medication or been in therapy for 8 weeks or longer. Daily benzodiazepine use was exclusionary. Participants taking benzodiazepines or opioids on an as-needed basis had to be willing to abstain from use within 24 hours of each float session. Other exclusion criteria included uncomfortability in water or receiving blood draws, any skin conditions or open wounds that could cause pain when exposed to saltwater, diagnosis of a neurological disorder (e.g., epilepsy), a comorbid diagnosis of bipolar disorder, schizophrenia, or an eating disorder, active suicidality with plan or intent, receiving current inpatient psychiatric treatment, or moderate to severe substance use disorder (as determined by the MINI 7.0). Evidence of current drug use was also assessed prior to each floatation-REST session via urine drug screen testing for amphetamines, methamphetamines, cocaine, barbiturates, benzodiazepines, methylenedioxymethamphetamine (MDMA), methadone,opiates, oxycodone, phencyclidine, propoxyphene, and ketamine, and breathalyzer testing for alcohol; a positive readout on any of these tests was exclusionary. Pregnancy was exclusionary based on a positive urine pregnancy test. All study procedures were approved by the local ethics committee (Western Institutional Review Board Protocol #: 20150528) and were performed in accordance with Declaration of Helsinki. All participants provided their written informed consent prior to participation and were compensated for their study involvement. The study was pre-registered on ClinicalTrials.gov (NCT03899090).

### Experimental Design and Procedures

Using a randomized parallel study design, participants completed a total of 11 visits. The study visit timeline is outlined in Figure 1. Participants initially completed a brief telephone screen to collect demographic information and evaluate potential study eligibility. During visit 1, official study eligibility was determined using the inclusion/exclusion criterion. After providing informed consent, participants returned for visit 2, where baseline symptom ratings were completed via clinician-reported and self-reported scales, and some behavioral testing of cardiac interoception (results to be reported separately). Visit 2 concluded by unblinding and informing the participant of their randomized (1:1:1) assignment to one of three intervention conditions: pool-REST, pool-REST preferred, or chair-REST (i.e., the active comparator, described further below). The randomization schedule was pre-determined by a statistical consultant uninvolved in the study before participant recruitment and stored as a blinded electronic list. At the time of assignment, the study coordinator would unblind and reveal the participant’s allocation. At the start of visit 3, all participants completed the Credibility/Expectancy Questionnaire (CEQ; [21]), to assess their pre-intervention attitudes towards the assigned intervention arm. Participants then completed six sessions (visits 3-8) according to their assigned condition. We assessed serious and non-serious adverse events after each session via a detailed survey of positive and negatively valenced experiences using a scale drawn from our prior floatation-REST study [5] and via verbal self-report throughout the study period. Following the sixth and final session (visit 8), participants returned to the lab for a post-intervention session (visit 9), where clinician-reported and self-reported symptom ratings were once again collected and behavioral testing of cardiac interoception was completed. The same clinician and self-reported symptom scales were completed again at 6-week (visit 10) and 6-month (visit 11) follow-up visits. Follow-up surveys were initially administered in the laboratory via an electronic tablet, but due to the Covid-19 pandemic these were switched to remote visits starting in May 2020. All surveys were obtained electronically using REDCap (Research Electronic Data Capture; Version 10.6.19; [22, 23]).

**Figure 1.**
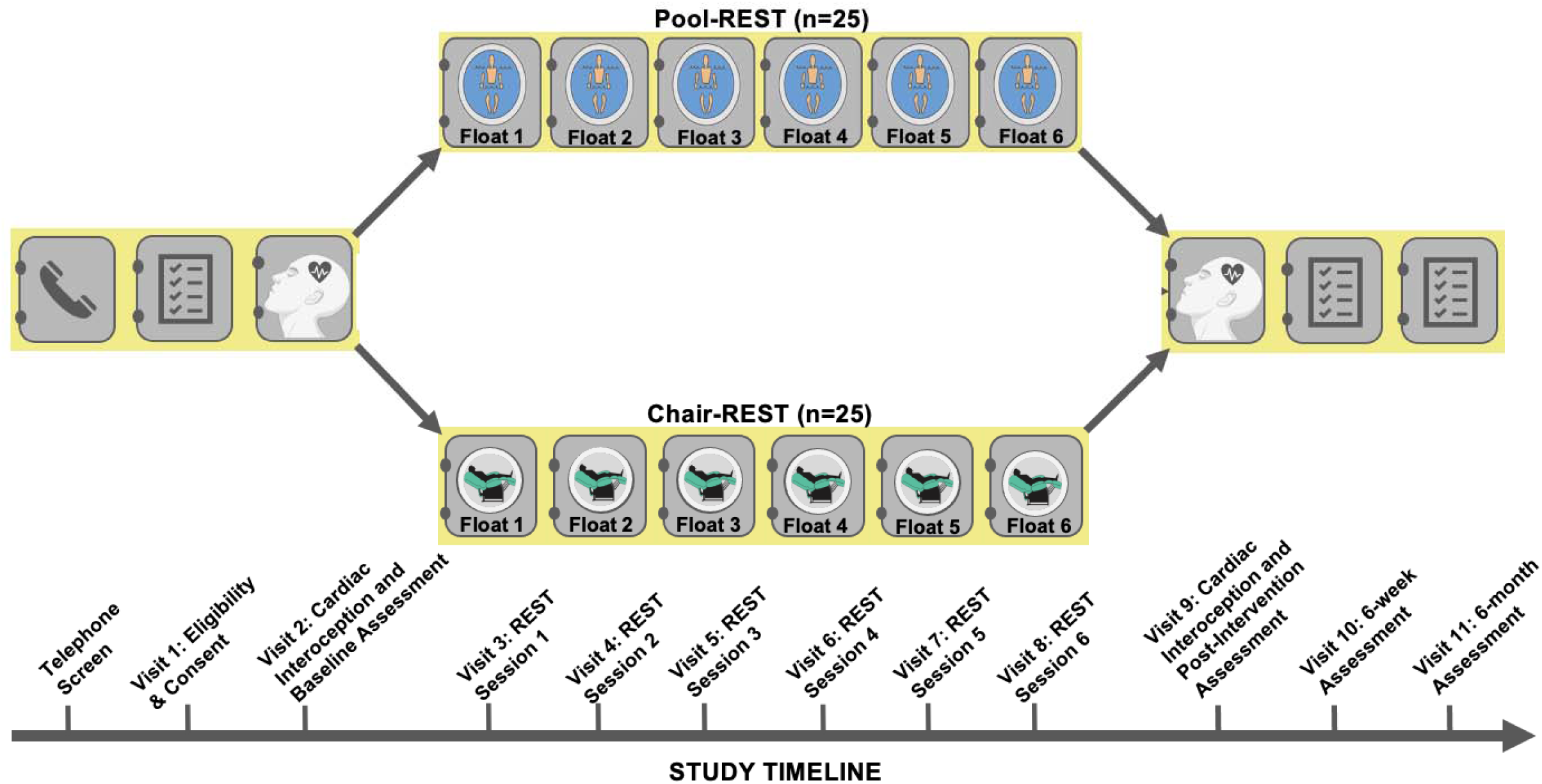
Study Visit Timeline.

### Experimental Conditions

#### Pool-REST

The pool-REST condition, which was delivered at the Laureate Institute for Brain Research facility, involved prescribed (i.e., fixed) session durations and inter-session intervals within an open or enclosed circular float pool (Floataway Inc., Norfolk, United Kingdom).

Participants were instructed to complete 1-hour float sessions once per week for six weeks. Each pool measured 2.44 meters in diameter and 0.28 meters in depth. The water in each pool was saturated with over 800 kilograms of Epsom salt (magnesium sulphate), creating a specific gravity of approximately 1.26. This dense salt-water solution allowed for participants to float effortlessly in a supine position. Both pools were constructed in rooms designed to be lightproof and soundproof. The air and water temperature of each room was also calibrated to match the temperature of the skin surface (∼35.0 degrees Celsius). More details about how the pool-REST condition was specially engineered to minimize all manner of external sensory stimulation can be found in Feinstein et al., 2018a. We allowed participants to choose between the open and enclosed pool to provide some flexibility for preference and to facilitate the optimal flow of participants through the study.

#### Pool-REST Preferred

The pool-REST preferred condition involved increased flexibility of both the session duration and the frequency. Participants were instructed that they were to complete six sessions in the open or closed pool, but they were allowed to freely set their float duration for up to two hours per session. In addition, they were permitted to choose the frequency of their floating schedule, such that they could float as often as they preferred within a 12-week period, with the only requirement being that there needed to be a minimum of 24 hours between sessions.

#### Chair-REST

During chair-REST, which was delivered at the Laureate Institute for Brain Research facility, participants reclined in a Zero Gravity Chair (PC510, Classic Power, Series 2, Human Touch Inc., Long Beach, CA). This active comparator condition was tailored to closely match the pool-REST condition on many parameters including a supine body position, the same prescribed session duration and inter-session interval (i.e., 1-hour sessions completed once per week for six weeks) in a room with reduced light and sound, and a similar instruction set emphasizing the importance of stillness and wakefulness throughout each session (see Supplemental Materials for full instruction set). Participants were informed that the chair was ergonomically designed to take pressure off the spinal cord and used memory foam backing to help the chair conform to each participant’s body shape. To match the instruction set across REST conditions, the Zero Gravity chair was explicitly referred to as the “float chair” and the act of lying in the chair was referred to as “floating.” The chair was placed in a dark and quiet room, although the degree of light and sound attenuation was not as strong as that delivered in the pool conditions (i.e., participants could perceive a low-level of external sound and light). Unlike the pool conditions, the ambient temperature was maintained at a normal room temperature of ∼23.3 degrees Celsius, so participants randomized to the chair-REST condition remained fully clothed during each session.

### Measures

#### Events Assessment

A 43-item self-report measure ([5]; see Supplemental Material) was used to assess safety and subjective experiences occurring in each float session. This checklist was adapted from our previous pilot studies and was designed to assess potential adverse experiences associated with floatation-REST. 29 of the 43 items assessed the presence of negative physical or psychological events including itchiness, dry mouth, pain, anxiety/panic, flashbacks, suicidality, and hallucinations, as well as the presence of positive experiences (e.g., heightened feelings of joy, refreshment, serenity); these 14 items were included to reduce response biases, as well as to prevent a sole focus on assessing negative experiences associated with REST. Each item was rated on four-point scale (“none,” “mild,” “moderate,” or “extreme”) using the following instructions: “*Did you notice or experience an increase in any of these items during or shortly after your float today? Please only mark items that showed an increase from your typical day-to- day experience*.” For any item that had an endorsement other than “None,” participants were provided a free-response box to describe their experience in more detail. For certain negative symptoms (strong emotional memories, flashbacks, heightened thoughts related to death, visual or auditory hallucinations, out-of-body experiences, feelings of detachment, loss of control over behavior, and flight of ideas/racing thoughts) receiving any endorsement other than “None” prompted participants with the following question: *“Overall, was this a positive or negative experience?”* [5]. Items rated as “none” were coded as 0, negatively experienced items ranged from -3 (“extremely negative”) to -1 (“mildly negative”), and positively experienced items ranged from 1 (“mildly positive) to 3 (“extremely positive”).

#### Credibility/Expectancy Assessment

The Credibility/Expectancy Questionnaire (CEQ; [21]) was used to measure attitudes about treatment feasibility. The CEQ is a 6-item self-report measure with individual items rated on a 1-9 or a 0%-100% scale. Items assess attitudes about “how believable, convincing, and logical a treatment is” (i.e., credibility; [24]), and expectations one holds about how the treatment might influence symptoms (i.e., expectancy).

There were no changes to the primary outcome or the main secondary outcomes reported in the current study. A number of additional secondary outcomes were specified in May, 2022, prior to analysis of the data, and will be included in a separate report.

## Statistical Analysis

### Feasibility

The primary outcome of feasibility was assessed by calculating adherence to the assigned sessions via the formula 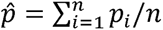, where *p_i_* was the proportion of completed sessions (out of 6) for subject and n was the number of subjects per condition (i.e., 25). To measure attitudinal aspects of feasibility, pre-treatment credibility beliefs were calculated by taking the mean of the first three items of the CEQ [25], while expectancy beliefs were captured via a single item rated on a 0 to 100% scale assessing how much improvement in their symptoms the participant thought would occur after completing the assigned intervention. Two separate one-way ANOVAs were performed to assess for between-group differences in pre-intervention credibility and expectancies, respectively.

### Tolerability

Tolerability was assessed in three ways; first, by calculating the overall dropout rate for each condition (i.e., (number of subjects who withdrew from the study or who were lost to follow-up)/25). Secondly, a Kaplan-Meier analysis with post-hoc log-rank tests was used to test for differential dropout rates over time for the three assigned conditions [26]. Third, within-session tolerability was assessed by fitting a linear mixed models (estimated using REML and the nloptwrap optimizer) to predict the duration of each session with visit and condition as fixed effects (formulas: Session Duration (in minutes) ∼ Visit * Condition). The model included subject ID as a random effect (formula: ∼1 | Subject ID) and utilized an AR1 covariance structure. Post-hoc two-sided t-tests with Holm corrections were used to interpret significant main effects, as well as simple effects for all significant interactions.

### Safety

The number of adverse and serious adverse events and their relationship to the intervention were recorded over the course of the entire study, including during the follow-up period. We then fit a linear mixed model (estimated using REML and the nloptwrap optimizer) to predict event magnitude ratings with event, visit, and condition parameters (formulas: Event Intensity Rating ∼ Event * Visit * Condition). The models included subject ID and visit as random effects (formula: ∼1 | Subject ID/Visit) and utilized an AR1 covariance structure. Post-hoc two-sided t-tests with Holm corrections (Holm, 1979) were used to interpret significant main effects, as well as simple effects for all significant interactions. All analyses were performed in RStudio (Version 3.6.0; [27]).

### Pool-REST Preferred

The impact of flexible assignment on preferred frequency of floating for the pool was measured by calculating the average number of days between float sessions and the corresponding standard deviation. To assess whether session frequency in this condition was related to baseline severity of anxiety, depression, and anxiety sensitivity symptoms, we examined bivariate correlations between float frequency and baseline OASIS, PHQ-9, and ASI-3 scores. Additionally, we examined bivariate correlations between the average session duration (in minutes) for each intervention and baseline measures of symptom severity (i.e., OASIS, PHQ-9, and ASI-3).

## Results

### Participant Characteristics

Recruitment occurred between February 2019 and October 2021, with a four month pause starting March 2020 during the Covid-19 pandemic and was terminated upon achievement of the recruitment target. Sociodemographic and clinical characteristics for the 75 individuals who were randomized to each intervention arm are summarized in Table 1. A CONSORT diagram reflecting the flow of participants in each arm is shown in Figure 2. One-way ANOVAs revealed that the groups did not differ significantly on age, education, medication status (i.e., number of psychotropic medications), BMI, or baseline OASIS, ASI-3, or PHQ-9 scores. Chi-squared tests indicated that there were no significant differences between the conditions regarding sex, race/ethnicity, or the frequency of clinical diagnosis or rate of psychotherapy utilization. Most of the sample identified as non-Hispanic White (81.3%). The most common psychiatric diagnosis in the sample was major depressive disorder (97.3%) followed by generalized anxiety disorder (50.7%).

**Figure 2.**
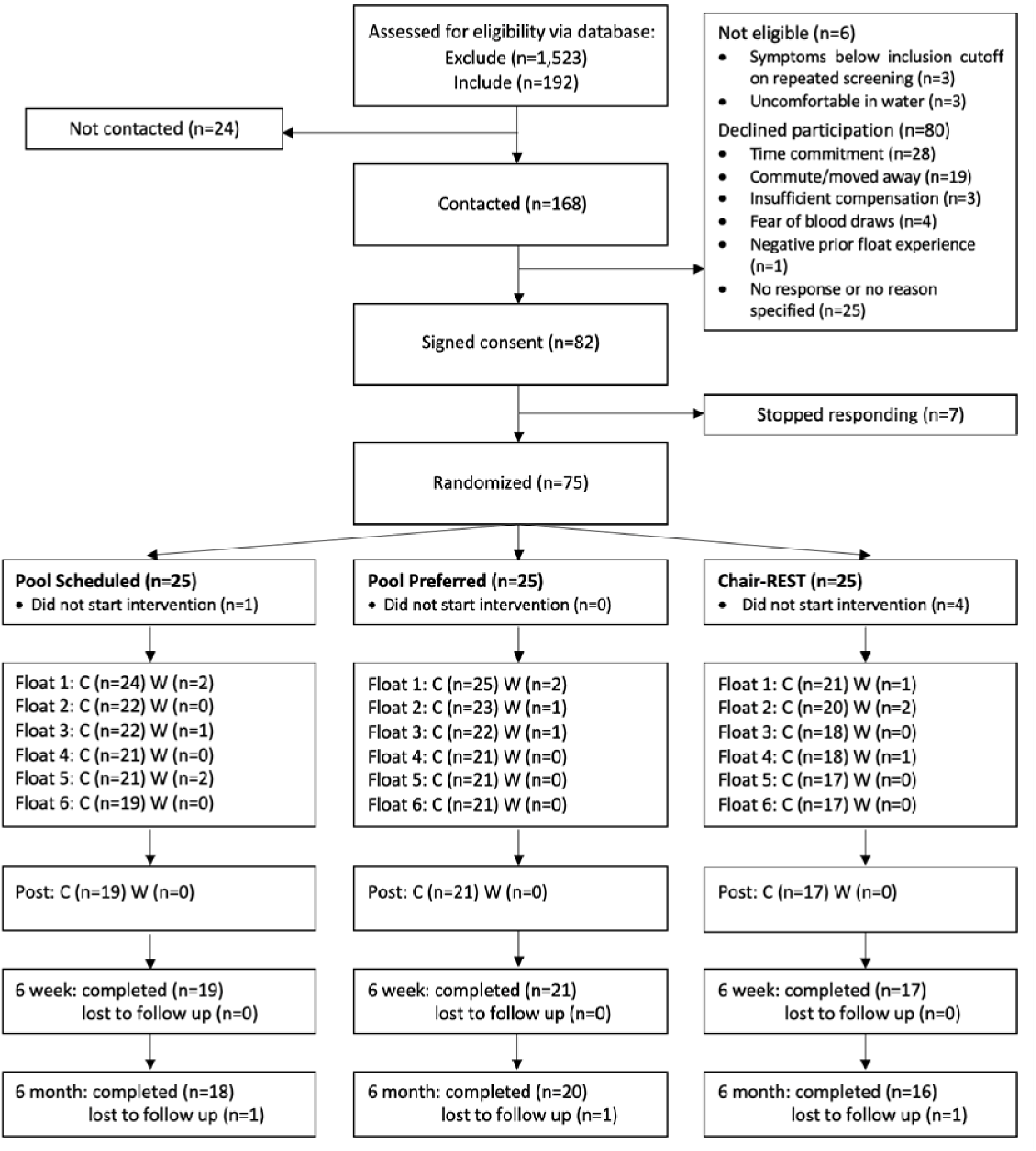
CONSORT Diagram. Note. C = Completed. W = Withdrew.

**Table 1.** Participant Demographics, DSM-5 Diagnostic Comorbidities, and Screening Scores at Study Entry

|  | *Chair-REST*<br>(n=25) | *Pool-REST*<br>(n=25) | *Pool-REST preferred*<br>(n=25) | *F* | *df* | *p* |
| --- | --- | --- | --- | --- | --- | --- |
| **Demographics** |  |  |  |  |  |  |
| Sex | | | | $\chi^2 = 0.15$ | 2 | 0.93 |
| Male, N (%) | 6 (24%) | 6 (24%) | 5 (20%) |  |  |  |
| Female, N (%) | 19 (76%) | 19 (76%) | 20 (80%) |  |  |  |
| Age | 37.8 (13.4) | 33.4 (11.7) | 34.3 (9.0) | 1.01 | 72 | 0.37 |
| Years of Education | 15.2 (1.3) | 15.1 (1.8) | 15.8 (1.7) | 1.15 | 71 | 0.32 |
| BMI | 29.3 (7.3) | 28.8 (5.1) | 27.9 (6.9) | 0.30 | 70 | 0.74 |
| Psychiatric Medications<sup>a</sup> | 0.7 (1.1) | 0.3 (0.6) | 1.0 (1.3) | 3.00 | 72 | 0.06 |
| Receiving Psychotherapy, N(%) | 11 (44) | 11 (44) | 16 (64) | $\chi^2 = 2.67$ | 2 | 0.26 |
| Race/Ethnicity, N(%) | | | | $\chi^2 = 8.75$ | 10 | 0.56 |
| White/Caucasian | 22 (88) | 20 (80) | 19 (76) |  |  |  |
| Black/African American | 2 (1) | 6 (24) | 4 (16) |  |  |  |
| Am. Indian/Alaskan Native | 5 (20) | 5 (20) | 4 (16) |  |  |  |
| Asian/Pacific Islander | 1 (4) | 0 (0) | 0 (0) |  |  |  |
| Hispanic | 1 (4) | 2 (8) | 2 (8) |  |  |  |
| Unspecified | 0 (0) | 0 (0) | 2 (8) |  |  |  |
| **Self-Report Measures** |  |  |  |  |  |  |
| OASIS Anxiety | 10.1 (2.5) | 9.4 (2.8) | 9.9 (2.5) | 0.53 | 69 | 0.59 |
| ASI-3 Anxiety Sensitivity | 35.6 (13.0) | 39.9 (9.4) | 40.5 (11.8) | 0.04 | 69 | 0.97 |
| PHQ-9 Depression | 12.5 (4.9) | 11.5 (5.5) | 12.1 (4.7) | 0.28 | 69 | 0.76 |
| **Psychiatric Diagnosis, N(%)<sup>b</sup>** | | | | $\chi^2 = 3.78$ | 8 | 0.88 |
| Major depressive disorder | 24 (96) | 24 (96) | 25 (100) |  |  |  |
| Generalized anxiety disorder | 10 (40) | 17 (68) | 11 (44) |  |  |  |
| Social anxiety disorder | 9 (36) | 12 (48) | 8 (32) |  |  |  |
| Posttraumatic stress disorder | 9 (36) | 8 (32) | 7 (28) |  |  |  |
| Panic disorder | 3 (12) | 6 (24) | 7 (28) |  |  |  |

*Note.* Numbers reflect means and standard deviation unless otherwise indicated. BMI = Body Mass Index; OASIS = Overall Anxiety Severity and Impairment Scale [18] measured at pre-intervention visit; ASI-3 = Anxiety Sensitivity Index-3 [19], measured at pre-intervention visit; PHQ-9 = Patient Health Questionnaire-9 [20] measured at pre-intervention visit. <sup>a</sup>Numbers reflect the mean number of prescribed psychiatric medications taken by the participant, measured at the initial screening. <sup>b</sup>Diagnosis was determined by clinician interview using the MINI version 7.0 [28]. For brevity, only comorbid diagnoses with frequency > 10% across the entire sample (n = 75) are listed. As this study allowed for comorbid diagnoses, percentage totals will be > 100%.

### Feasibility

Average treatment adherence was 89% (5.3 1.6 sessions) for the pool-REST preferred condition, 85% (5.1 1.8 sessions) for the pool-REST condition, and 74% (4.4 2.5 sessions) for the chair-REST condition. One-way ANOVAs revealed that the REST conditions did not differ significantly on pre-intervention CEQ credibility (F(2, 67) = 2.42, *p* = 0.097) or expectancy (F(2, 67) = 0.98, *p* = 0.379; see Figures 3a-3b). The average credibility score for the three groups was 6.67 ± 1.50. On average, the three groups expected the intervention to halve their symptoms (50.3 ± 21.6%).

**Figure 3.**
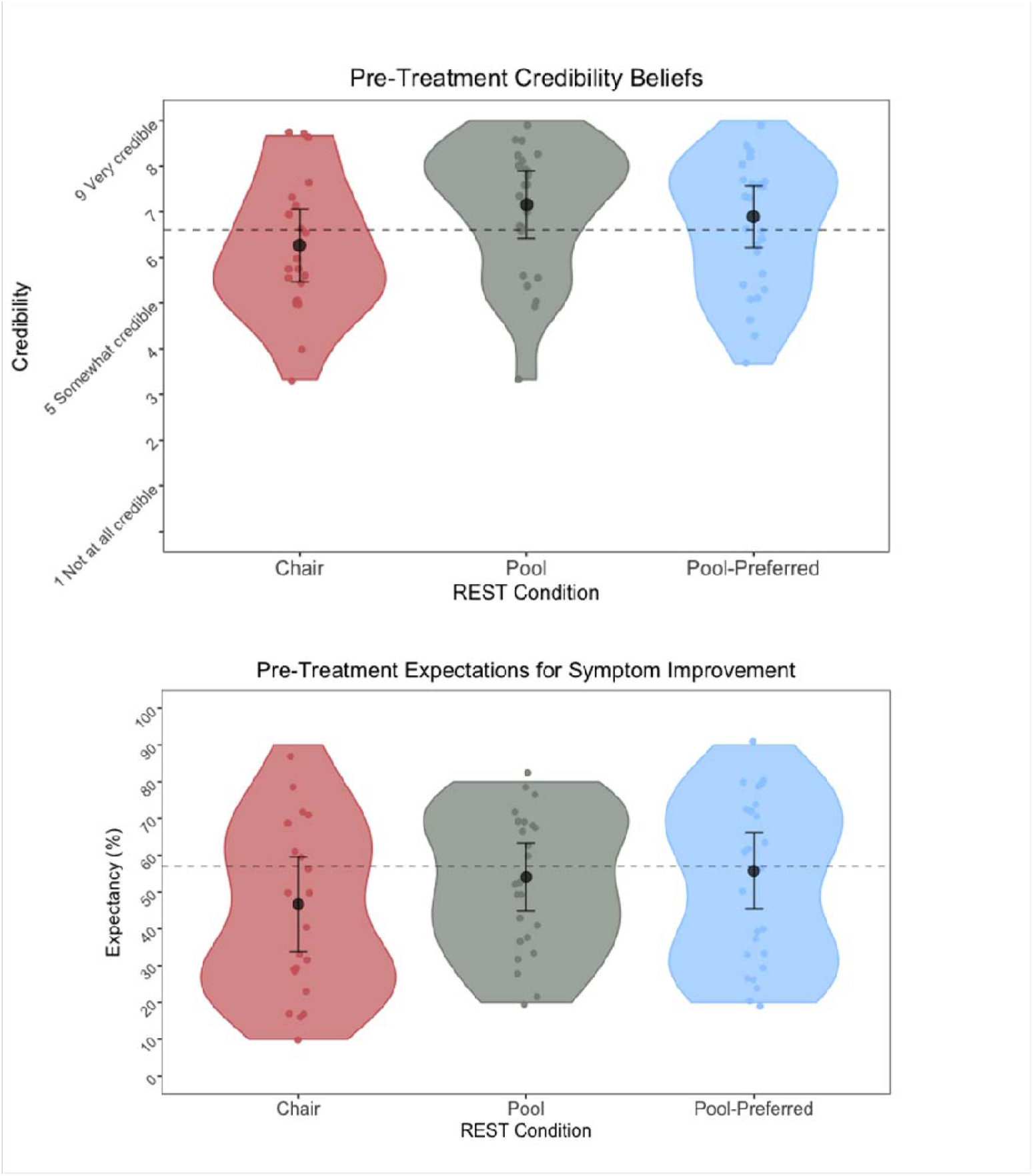
Pre-Treatment Credibility Beliefs and Expectations for Symptom Improvement. *Note.* Figures reflect pre-intervention ratings for credibility of beliefs (top) and expectations for symptom improvement (bottom) on the Credibility and Expectancy Questionnaire, according to the assigned intervention. The black dots and error bars represent the mean and standard error of the mean, respectively. Grey dashed lines reflect an established average for credibility and expectancy beliefs of other non-pharmacological treatments (e.g., psychotherapy, see discussion). There were no significant group differences in levels of pre-treatment credibility or expectancy beliefs (*p*s > .05).

### Tolerability

The Kaplan-Meier survival analyses did not indicate a significant difference in treatment dropout among the three groups (*p* = .40, Figure 4). Following the randomization at the baseline visit, the chair-REST condition had the highest dropout rate at 16% (n = 4 individuals), versus 4% (n = 1 individual) for the pool-REST condition and 0% for the pool-REST preferred condition. Across the six-session intervention, the chair-REST condition demonstrated an overall dropout rate of 32%, while the pool-REST condition dropout rate was 24%, and the pool-REST preferred dropout rate was 16%. The dropout rate at the 6-month follow up was 36% for the chair-REST condition, 28% for the pool-REST condition, and 20% for the pool-REST preferred condition (1 individual dropped out from each of the groups).

**Figure 4.**
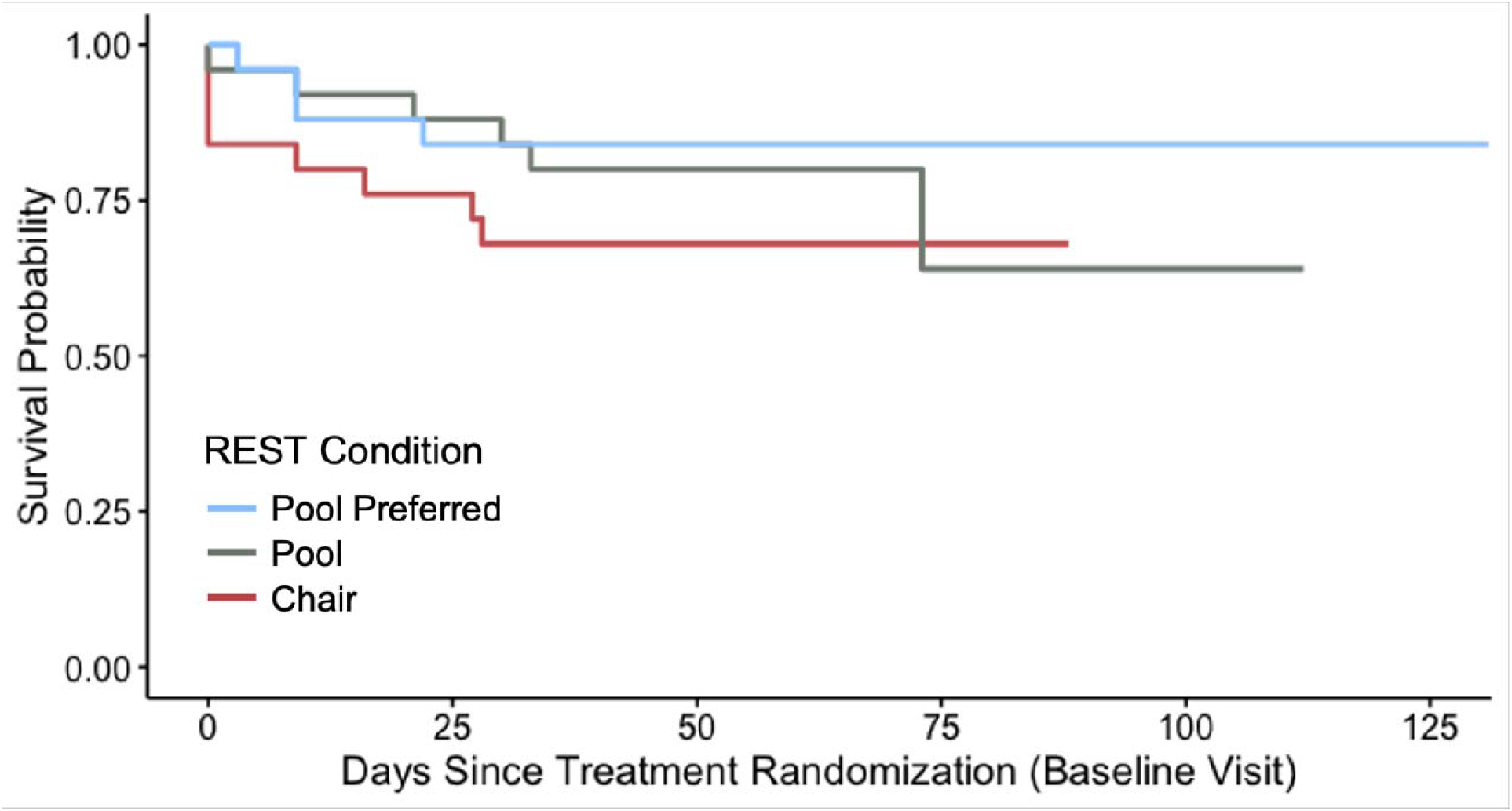
Kaplan-Meier Survival Curve illustrating dropout over time for each REST Condition.

Of note, due to the onset of the Covid-19 pandemic and mandatory shutdowns, two participants were withdrawn from the study by the study investigators. One participant in the Chair-REST condition was withdrawn from the study after session 2 (at day 27), and one participant from the Pool-REST condition was withdrawn following their fifth session (at day 73). This dropout information was entered into the analysis, but due to the small amount of dropout and no discernable group pattern, no further corrective action was taken during the analysis.

The linear mixed effects analysis of within-session tolerability (i.e., session duration; Figure 5), revealed a main effect of condition (F(2) = 10.33, *p* < .001, h ^2^ = .24), such that session durations were significantly longer for the pool-REST preferred condition (*M* = 75.4 ± 29.4 minutes) than either the chair-REST (*M* = 58.4 ± 4.3 minutes; *p* = .007, *d* = .810) or pool- REST condition (*M* = 53.0 ± 12.3 minutes; *p* = .0002, *d* = .996). There were no statistically significant differences in session duration between the chair-REST and pool-REST conditions (*p* = .292). Across the 373 REST sessions administered across the entire study, 317 sessions (85% of total) were 50 minutes or longer in duration.

**Figure 5.**
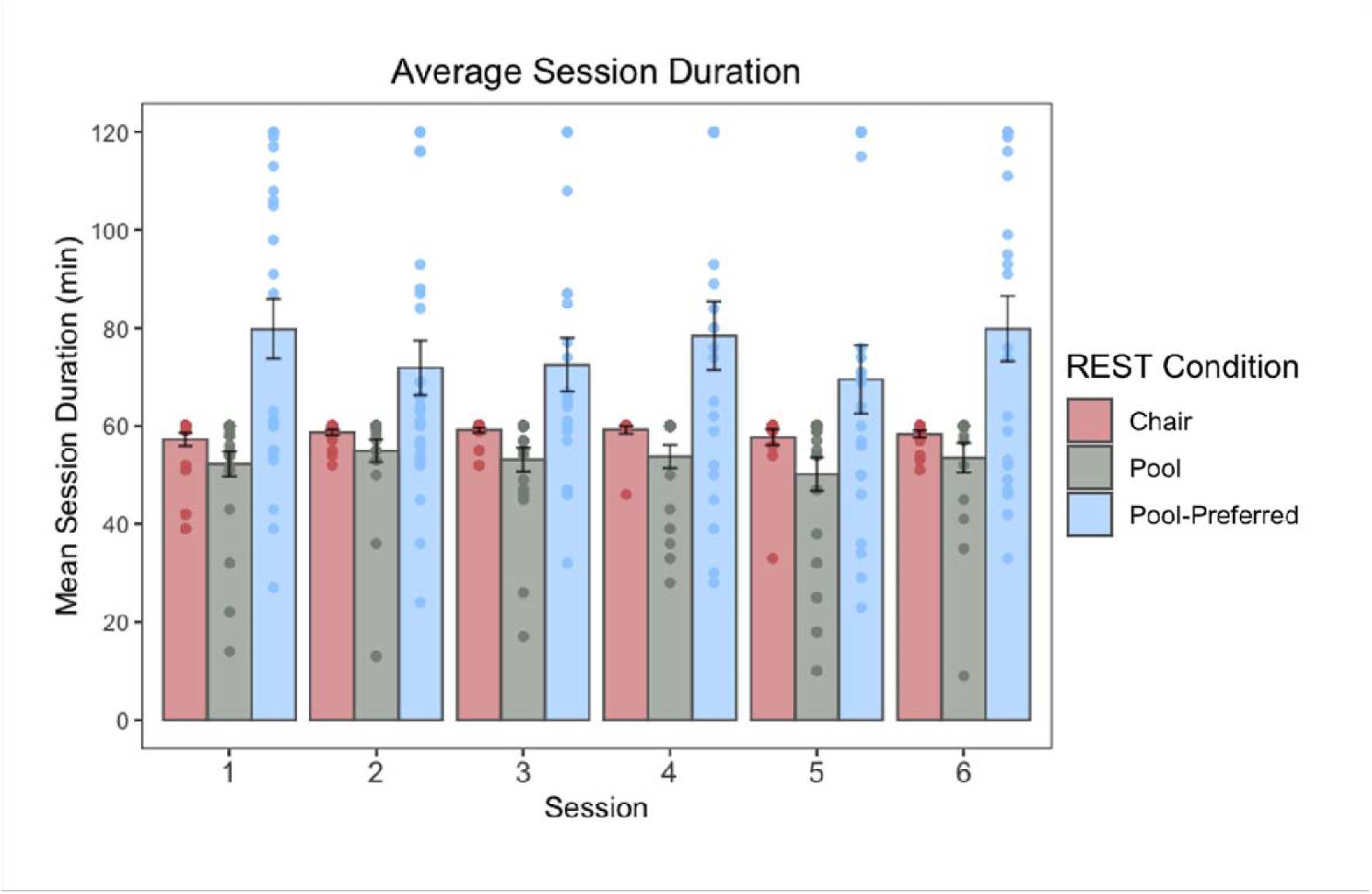
Session durations. Note. Session durations were significantly longer for the pool- REST preferred condition than either the chair-REST (p = .007) or pool-REST conditions (p = .0002). There were no statistically significant differences in session duration between the chair- REST and pool-REST conditions (p = .292). Duration did not vary as a function of session.

### Safety

Four adverse events were recorded during the study. All were determined to be unrelated to the study intervention. One participant in the chair-REST condition reported two separate instances of migraines with nausea on the days of their first and second REST session. One participant in the pool-REST condition who suffered from seasonal allergies reported an ear infection four days after their third REST session. Finally, one participant in the pool-REST condition reported a suicide attempt the night before their 6-week follow-up visit. The participant was subsequently evaluated at a local psychiatric emergency department, where it was determined that hospital admission was unwarranted.

Regarding the acutely experienced event profile, Figure 6 shows the average magnitude rating for each event across the three assigned REST conditions. A qualitative inspection of the data revealed that positive experiences were endorsed more commonly than negative ones and were also rated at higher levels of intensity, with average intensity ratings in the mild-to-moderate range for positive experiences and well-below mild for negative experiences. In the linear mixed effects (LME) quantitative analysis of intensity ratings, we observed a significant main effect of event (F(43) = 153.61, *p* < .001, h ^2^ = .30) and treatment condition (F(2) = 3.51, *p* = .04, h ^2^ = .09), which was accounted for by the significant interaction between these variables (F(86) = 6.43, *p* < .0001, h ^2^ = .04). Post-hoc comparisons suggested that feelings of intense joy/happiness, increased energy, increased ability to focus/concentrate, serenity and peacefulness, appreciation for life, refreshment, relaxation, silent mind, pain-free existence, and feelings of flow were experienced more positively in the pool-REST and pool-REST preferred conditions when compared to the chair-REST condition (*p*s < .05). Stronger positive ratings for the pool-REST preferred condition were also observed for relaxation and pain-free existence when compared to the pool-REST condition (*p*s < .05). The opposite trend was found for feelings of flow, increased empathy and compassion for others, and joy/happiness, such that participants in the pool-REST condition tended to rate these items more positively than their pool-REST preferred counterparts (*p*s < .05). The pool-REST group also rated feelings of empathy and compassion for others more positively than their chair-REST counterparts (*p* < .01).

**Figure 6.**
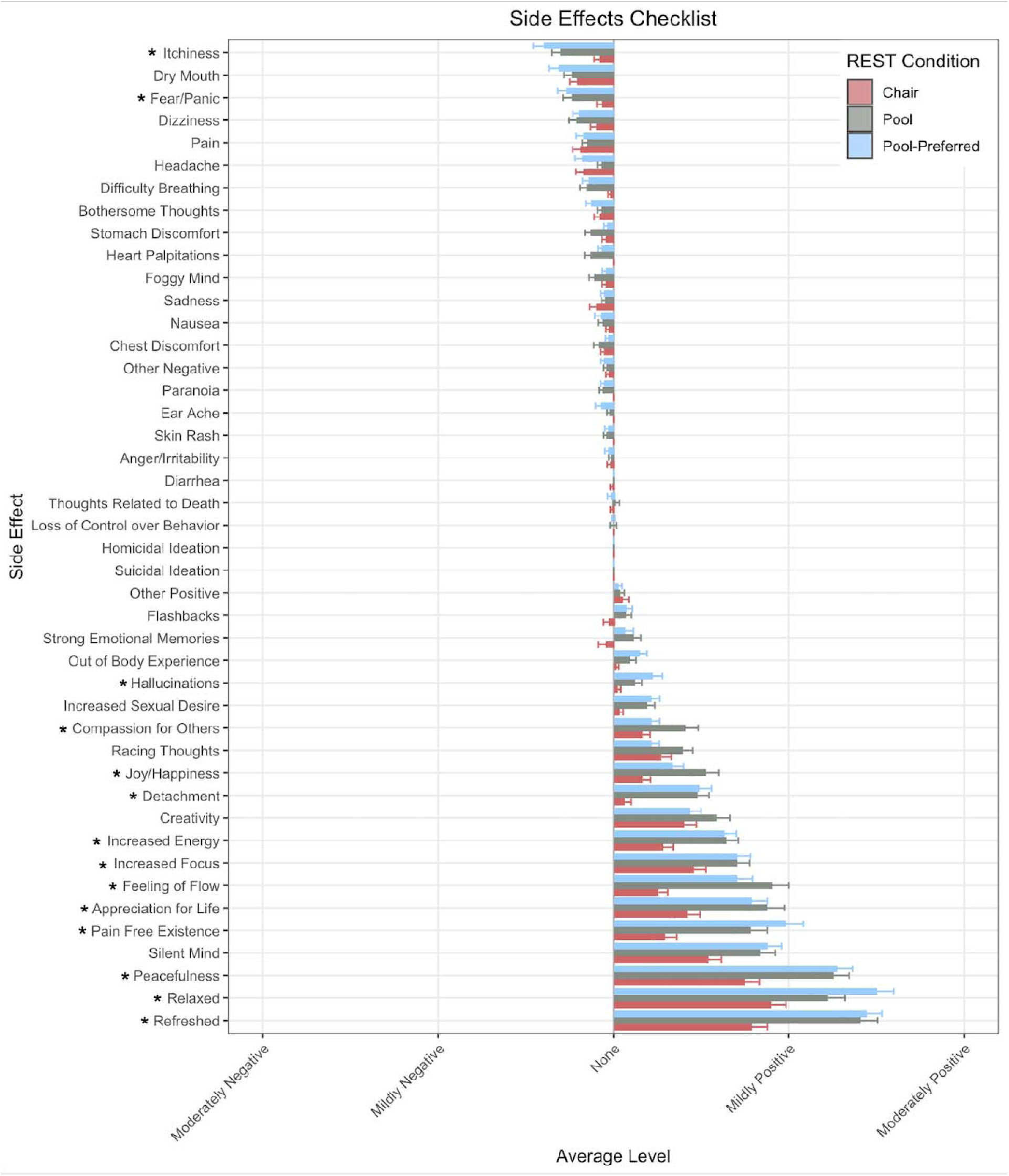
Events Checklist. Note. Event anchors ranged from “Extremely Negative” to “Extremely Positive” (anchors not shown). Asterisk (*) indicates significant group difference in rating magnitude.

Regarding events that typically carry negative connotations, post-hoc comparisons suggested that itchiness was experienced more negatively in the pool-REST and pool-REST preferred conditions than the chair-REST condition (*p*s < .05). The pool-REST preferred condition also rated feelings of intense fear and panic more negatively than the chair-REST condition (*p* = .045). Interestingly, the pool-REST and pool-REST preferred conditions endorsed experiencing feelings of detachment more positively than the chair-REST condition (*p*s < .001).

Stronger positive ratings were also observed in the pool-REST preferred condition than the chair-REST condition for hallucinations (*p* = .03). Of note, the reports of hallucinations and feelings of detachment occurred infrequently and tended to be described in neutral or positive terms. For example, descriptions of hallucinations commonly mentioned visually experienced colors and shapes, such as “*Colorful visions; shapes, people, and characters*,” “*Saw green, crystal-like geometric shapes off and on (eyes were closed)*,” and “*I experienced those colorful flashes and dots similar to when you close your eyes and press lightly on your eyes.*” Feelings of detachment were typically described using feelings of relaxation, weightlessness, and disconnection from space and time, including: “*I felt weightless. I felt like nothing else mattered. It was just me*,” “*Felt very relaxed like everything was far away*,” “*A similar dozing detachment as when you’re half asleep for a really nice nap*,” and “*I felt like I was in my own world and the real world was frozen in time.*”

The linear mixed model analysis also revealed a significant interaction between event and visit (F(214) = 1.40, *p* = .0001, h ^2^ = .02). Specifically, post-hoc analyses showed a weakening in the magnitude of positive events for all three interventions over the course of the study for the following events: serenity and peacefulness, appreciation for life, refreshment, relaxation, silent mind, pain-free existence, and feelings of flow (*p*s < .05).

### Pool-REST Preferred

When allowed to select the frequency of sessions, the pool-REST preferred participants chose to float at a frequency of once every 12 days, on average (*SD =* 9.33), and for an average float duration of 75 minutes (*M* = 75.4 ± 29.4 minutes). Exploratory analyses indicated that session frequency and session duration were not significantly associated with baseline measures of anxiety, depression, or anxiety sensitivity (*p*s > .05).

## Discussion

This study represents the first randomized controlled trial examining the feasibility, tolerability, and safety of repeated sessions of floatation-REST in individuals with anxiety and depression. We observed evidence of a favorable feasibility, tolerability, and safety profile for the application of floatation-REST and chair-REST in this outpatient sample. Positive experiences were endorsed more commonly than negative ones and were also rated at higher levels of intensity, with no serious adverse events associated with any intervention. Taken together, six sessions of floatation-REST appear feasible, well-tolerated, and safe in anxious and depressed outpatient individuals.

Our primary outcome of adherence suggested that the feasibility of each form of REST, indexed via adherence to the allocated six-sessions, was adequate, ranging from 85-89% for the pool-REST conditions to 74% for the chair-REST condition. These levels of adherence are comparable to traditional behavioral interventions for anxiety and depression, whose treatment completion rates hover around 80% [16]. Each REST intervention was perceived to be moderately credible, with participants having positive expectations for symptom improvement.

The observed levels of credibility and expectancy beliefs are also consistent with those typically seen for other behavioral interventions such as exposure therapy (range of credibility beliefs = 5.95-7.47, range of expectancy beliefs = 50.50-67.90; [21, 25, 29]).

The low dropout rates in this behavioral intervention study provides additional evidence that repeated sessions of REST are well-tolerated. Dropout rates in the post randomization setting prior to intervention onset were lowest for the pool interventions (4% dropout for pool- REST, and 0% dropout for pool-REST preferred), while the chair-REST pretreatment dropout rate was the highest (16%) and similar to those observed to cognitive behavioral therapy (dropout rates ranging from 11-21%, [15]). Dropout rates during the administration of the REST intervention (32% for chair-REST, 24% for pool-REST, and 16% for pool-REST preferred) were also comparable to rates seen in non-pharmacological and pharmacological interventions such as cognitive behavioral therapy (19-36%, [15]), meditation (33-44%, [30]), acceptance and commitment therapy (15%, [31]), and antidepressant medications (37-49%, [32]).

Using session duration as an additional proxy of intervention tolerability, we saw very few individuals in the pool-REST and chair-REST conditions who had a prescribed session duration (i.e., 60 minutes) terminate early, with the majority of individuals utilizing their full time allotment. For the pool-REST preferred group, who was given greater flexibility in session duration (i.e., allowed to float for up to two hours), we observed longer average session durations. Although the preferred condition was given an additional 60 minutes, they tended to float about 20 minutes longer than the prescribed session conditions on average. Moreover, session duration was unrelated to symptom severity at baseline, indicating that individuals with higher levels of anxiety, depression, or anxiety sensitivity were no more or less likely to utilize longer durations (i.e., in a self-medicating or symptom-dependent fashion). Taken together, these findings may suggest that a 60-minute float is a suitable duration for future clinical trials in a clinically anxious and depressed sample. However, it remains to be determined whether there is additional utility of more time (e.g., 75 minutes), or alternatively, whether similar effects could be achieved with a shorter durations (e.g., 30 or 45 minutes).

The safety findings from this trial are consistent with previous single-session studies of pool-REST and chair-REST which found few negative events and no serious adverse events [5, 7, 14]. Our study expands upon this work by showing a favorable safety profile across repeated sessions of floatation-REST. We also demonstrated that positive experiences were endorsed more commonly than negative ones, and at higher levels of intensity, in the mild-to-moderate range. Importantly, while all three intervention groups consistently rated positive effects of the intervention, the pool-REST groups tended to rate positive events more strongly, especially regarding feelings of peace and tranquility. These findings provide support for the overall safety of floatation-REST delivered in a pool or in a Zero Gravity chair setting. Moreover, the comparable intensity ratings of many negative events but weaker ratings of positive events within the chair-REST condition suggests that the chair is a suitable active comparator.

There were differential event profiles between the pool-REST conditions and the chair- REST condition, such that participants in the pool conditions reported stronger feelings of tranquility (e.g., refreshment, peacefulness, serenity) than those in the chair-REST condition and tended to rate itchiness and fear/panic more negatively than their chair-REST counterparts. The most common negative events associated with pool-REST were itchiness and dry mouth, which is consistent with our previous pilot study [5], and may be related to the high salt concentration in the pools. In addition, several experiences that traditionally carry negative connotations, including audiovisual hallucinations and feelings of detachment, tended to be described in a positive light in the pool conditions. We do not believe the hallucinations reported by participants in this study reflect hallucinations in the true sense of the term and do not consider them indicative of psychosis-spectrum symptoms. Rather, these visual and auditory sensations may be the result of the brain ‘filling in the gaps,’ i.e., generating sensory representations in the face of absent exteroceptive input to the nervous system. Finally, the repeated nature of this intervention allowed for a thorough examination of event profiles over time. An attenuation in the magnitude of several positive events was observed at later sessions for all three interventions, raising the possibility that the increased familiarity may reduce the novelty of the intervention and the magnitude of positive events.

This investigation had several limitations that must be acknowledged. First, four individuals in the chair-REST condition and one participant in the pool-REST condition withdrew from the study after randomized allocation to their respective intervention and did not complete the CEQ. These dropouts may suggest the presence of unmet expectations, particularly for the individuals allocated to the chair-REST condition given that the largest dropout occurred in this group. Alternatively, pre-intervention anticipatory anxiety could have played a role in withdrawal, although we did not receive such reports from participants. To avoid the limitation of missing data in the future, subsequent studies should aim to collect the CEQ measure immediately following randomization. Another limitation of the current study is the modest sample size. While our sample size was consistent with those used in previous floatation-REST trials, larger groups may be needed for an adequately powered survival analysis for the purposes of tolerability assessment. As such, the current study may be underpowered to detect meaningful group differences in dropout rate. We examined six-session feasibility of REST whereas prior clinical trials have typically used more sessions (e.g., 12). Thus, the measurement of feasibility and tolerability during the employment of a larger number of sessions may be advisable in future trials. Additionally, most of our sample was comprised of Caucasian, highly educated females, thereby limiting generalizability claims to the broader population. Finally, due to the fact that participants were financially compensated for their participation, we did not examine structural aspects of feasibility relevant to the real-world delivery of behavioral therapies, including financial and time commitments that might otherwise be important for treatment engagement.

## Conclusion

The findings from this clinical trial suggests that six sessions of REST in a chair or pool environment are feasible, well-tolerated, and safe in clinically anxious and depressed outpatient individuals. Larger randomized controlled trials evaluating markers of clinical efficacy are warranted.

## Data Availability

All data produced in the present study are available upon reasonable request to the authors.

## Funding

This research was supported by an award from the National Center for Complimentary and Integrative Health (R34AT009889). The authors were also supported by the National Institute of Mental Health (R01MH127225, K23MH112949 to SSK), National Institute of General Medical Sciences Center Grant Award Number (1P20GM121312 to MPP, JSF, SSK), and The William K. Warren Foundation. The funding sources had no role in the design and conduct of the study; collection, management, analysis, and interpretation of the data; preparation, review, or approval of the manuscript; or decision to submit the manuscript for publication.

## Author Contributions

McKenna Garland: Writing—Original draft preparation, investigation, formal analysis, visualization. Raminta Wilson: Investigation. Wesley Thompson: Writing—Review and editing. Murray Stein: Writing—Review and editing. Martin Paulus: Writing—Review and editing, supervision. Justin Feinstein: Conceptualization, methodology, resources, writing—review and editing, supervision, project administration, funding acquisition. Sahib Khalsa: Conceptualization, methodology, resources, writing—review and editing, supervision, project administration, funding acquisition.

## Declaration of Conflicting Interests

The authors declare that there are no conflicts of interests.

## Acknowledgments

We would like to thank Henry Yeh, Jamie Rhudy, Emily Adamic, and Joel Greenshields for consultation on coding and statistical matters. We would also like to acknowledge Laura Garrison for her contributions to data collection. Finally, we would like to thank the custodial staff at the Laureate Institute for Brain Research for their diligence in maintaining the floatation- REST and chair-REST facilities.

## Notes

### Competing Interest Statement

The authors have declared no competing interest.

### Clinical Trial

ClinicalTrials.gov NCT03899090

### Clinical Protocols

https://clinicaltrials.gov/ct2/show/NCT03899090

### Author Declarations

All study procedures were approved by the Western Institutional Review Board (Protocol #: 20150528) and were performed in accordance with Declaration of Helsinki. All participants provided their written informed consent prior to participation and were compensated for their study involvement.

### Summary of Updates

A sentence fragment was removed from the abstract.

## References

1. Suedfeld P, Coren S. Perceptual isolation, sensory deprivation, and rest: Moving introductory psychology texts out of the 1950s. Canadian Psychology/Psychologie canadienne. 1989;30(1):17.

2. Rossi AM. General methodological considerations. Sensory deprivation: Fifteen years of research. 1969:16–46.

3. Lilly JC, Shurley JT. Experiments in solitude, in maximum achievable physical isolation with water suspension, of intact healthy persons. Psychophysiological aspects of space flight. 1961:238–47.

4. Shurley JT. Profound experimental sensory isolation. American Journal of Psychiatry. 1960;117(6):539–45.

5. Feinstein JS, Khalsa SS, Yeh HW, Wohlrab C, Simmons WK, Stein MB, et al. Examining the short-term anxiolytic and antidepressant effect of Floatation-REST. PLoS One. 2018;13(2):e0190292. Epub 20180202. doi: 10.1371/journal.pone.0190292. PubMed PMID: 29394251; PubMed Central PMCID: PMCPMC5796691.

6. Al Zoubi O, Misaki M, Bodurka J, Kuplicki R, Wohlrab C, Schoenhals WA, et al. Taking the body off the mind: Decreased functional connectivity between somatomotor and default-mode networks following Floatation-REST. Hum Brain Mapp. 2021;42(10):3216–27. Epub 20210409. doi: 10.1002/hbm.25429. PubMed PMID: 33835628; PubMed Central PMCID: PMCPMC8193533.

7. Khalsa SS, Moseman SE, Yeh H-W, Upshaw V, Persac B, Breese E, et al. Reduced environmental stimulation in anorexia nervosa: an early-phase clinical trial. Frontiers in psychology. 2020:2534.

8. Solutions FT, LLC FO. State of the Float Industry Report. Float Tank Solutions; 2020.

9. Jonsson K, Kjellgren A. Promising effects of treatment with flotation-REST (restricted environmental stimulation technique) as an intervention for generalized anxiety disorder (GAD): a randomized controlled pilot trial. BMC complementary and alternative medicine. 2016;16(1):1–12.

10. Bood SÅ, Sundequist U, Kjellgren A, Nordström G, Norlander T. Effects of flotation rest (restricted environmental stimulation technique) on stress related muscle pain: are 33 flotation sessions more effective than 12 sessions? Social Behavior and Personality: an international journal. 2007;35(2):143–56.

11. Feinstein JS, Khalsa SS, Yeh H, Al Zoubi O, Arevian AC, Wohlrab C, et al. The elicitation of relaxation and interoceptive awareness using floatation therapy in individuals with high anxiety sensitivity. Biological psychiatry: cognitive neuroscience and neuroimaging. 2018;3(6):555–62.

12. Lann MA, Martin A. An unusual death involving a sensory deprivation tank. Journal of forensic sciences. 2010;55(6):1638–40.

13. Norlander T, Kjellgren A, Archer T. The experience of flotation-REST as a function of setting and previous experience of altered state of consciousness. Imagination, Cognition and Personality. 2000;20(2):161–78.

14. Forgays DG, Belinson MJ. Is flotation isolation a relaxing environment? Journal of Environmental Psychology. 1986;6(1):19–34.

15. Fernandez E, Salem D, Swift JK, Ramtahal N. Meta-analysis of dropout from cognitive behavioral therapy: Magnitude, timing, and moderators. Journal of Consulting and Clinical Psychology. 2015;83(6):1108.

16. Swift JK, Greenberg RP, Tompkins KA, Parkin SR. Treatment refusal and premature termination in psychotherapy, pharmacotherapy, and their combination: A meta-analysis of head- to-head comparisons. Psychotherapy. 2017;54(1):47.

17. Victor TA, Khalsa SS, Simmons WK, Feinstein JS, Savitz J, Aupperle RL, et al. Tulsa 1000: a naturalistic study protocol for multilevel assessment and outcome prediction in a large psychiatric sample. BMJ open. 2018;8(1):e016620.

18. Campbell-Sills L, Norman SB, Craske MG, Sullivan G, Lang AJ, Chavira DA, et al. Validation of a brief measure of anxiety-related severity and impairment: the Overall Anxiety Severity and Impairment Scale (OASIS). Journal of affective disorders. 2009;112(1-3):92–101.

19. Taylor S, Zvolensky MJ, Cox BJ, Deacon B, Heimberg RG, Ledley DR, et al. Robust dimensions of anxiety sensitivity: development and initial validation of the Anxiety Sensitivity Index-3. Psychological assessment. 2007;19(2):176.

20. Kroenke K, Spitzer RL, Williams JB. The PHQLJ9: validity of a brief depression severity measure. Journal of general internal medicine. 2001;16(9):606–13.

21. Devilly GJ, Borkovec TD. Psychometric properties of the credibility/expectancy questionnaire. Journal of behavior therapy and experimental psychiatry. 2000;31(2):73–86.

22. Harris PA, Taylor R, Thielke R, Payne J, Gonzalez N, Conde JG. Research electronic data capture (REDCap)—a metadata-driven methodology and workflow process for providing translational research informatics support. Journal of biomedical informatics. 2009;42(2):377–81.

23. Harris PA, Taylor R, Minor BL, Elliott V, Fernandez M, O’Neal L, et al. The REDCap consortium: Building an international community of software platform partners. Journal of biomedical informatics. 2019;95:103208.

24. Kazdin AE. Therapy outcome questions requiring control of credibility and treatment- generated expectancies. Behavior Therapy. 1979;10(1):81–93.

25. Thompson-Hollands J, Bentley KH, Gallagher MW, Boswell JF, Barlow DH. Credibility and outcome expectancy in the unified protocol: Relationship to outcomes. Journal of Experimental Psychopathology. 2014;5(1):72–82.

26. Kaplan EL, Meier P. Nonparametric estimation from incomplete observations. Journal of the American statistical association. 1958;53(282):457–81.

27. Team R. RStudio: Integrated Devleopment for R. . Boston, MA: RStudio, PBC; 2020.

28. Sheehan D, Janavs J, Baker R, Harnett-Sheehan K, Knapp E, Sheehan M. Mini international neuropsychiatric interview. Tampa: University of South Florida. 1994.

29. Arch JJ, Twohig MP, Deacon BJ, Landy LN, Bluett EJ. The credibility of exposure therapy: Does the theoretical rationale matter? Behaviour research and therapy. 2015;72:81–92.

30. Krisanaprakornkit T, Sriraj W, Piyavhatkul N, Laopaiboon M. Meditation therapy for anxiety disorders. Cochrane Database of Systematic Reviews. 2006;(1).

31. Ong CW, Lee EB, Twohig MP. A meta-analysis of dropout rates in acceptance and commitment therapy. Behaviour research and therapy. 2018;104:14–33.

32. Machado M, Iskedjian M, Ruiz I, Einarson TR. Remission, dropouts, and adverse drug reaction rates in major depressive disorder: a meta-analysis of head-to-head trials. Current medical research and opinion. 2006;22(9):1825–37.

