## Supplemental Information for "A randomized controlled safety and feasibility trial of floatation-REST in anxious and depressed individuals"

**Supplemental Materials**

***Sample Size Determination***

No power analyses were conducted for this early-phase trial, following the guidance of the funding opportunity announcement (FOA) associated with the funded grant supporting this study. Sample size was determined by assuming the distribution of individual adherence proportion is approximately symmetric, with a sample size of 25 subjects per group providing an adherence estimate with margin of error: t_0.975,24_/$\surd25$= 0.41 times the standard deviation at 95% confidence. Given the paucity of previous float research in this target population, it was also difficult to accurately estimate the number of individuals who would drop out of the study. We used rates of dropout for psychotherapy studies for anxiety and depression, which have an estimated attrition rate of ~20% (Taylor et al., 2012; Hembree et al., 2003) as a proxy, which would equate to a total of 60 completers (20 participants/condition). Therefore, we estimated that 25 participants per condition should be sufficient for examining basic patterns of adherence.

***Events Checklist***

**Instructions**: Did you notice or experience an INCREASE in any of the items below during or shortly after your float today? Please only mark items that showed an increase from your typical day-to-day experience.

**Items**:

1. Heightened creativity
2. Flight of ideas or racing thoughts
   a. Overall, was this a positive or negative experience?
3. Feelings of intense euphoria, joy, or happiness
4. Heightened energy
5. Heightened focus and ability to concentrate
6. A feeling of total serenity and peacefulness
7. Increased sexual desire
8. Heightened empathy and compassion for others
9. A strong feeling of appreciation that you are alive
10. Feeling completely refreshed, like the reset button was hit
11. Total relaxation of the body (without any muscle tension)
12. Complete silence of mind (without any anxious thoughts or worries)
13. Totally pain-free existence
14. A feeling of “flow” with the world around you
15. Dizziness
16. Headache or migraine
17. Feelings of intense fear, anxiety, or panic
18. Difficulty breathing
19. Heart palpitations
20. Chest discomfort
21. Stomach discomfort
22. Nausea
23. Diarrhea
24. Pain
25. Itchiness
26. Skin rash
27. Dry mouth
28. Ear ache
29. Strong emotional memories
    a. Overall, was this a positive or negative experience?
30. Flashbacks (reliving a memory as if it were happening all over again)
    a. Overall, was this a positive or negative experience?
31. Heightened thoughts related to death
    a. Overall, was this a positive or negative experience?
32. A desire or wanting to hurt or kill yourself
33. A desire or wanting to hurt or kill others
34. Visual or auditory hallucinations
    a. Overall, was this a positive or negative experience?
35. Paranoia (intense fear or suspicion of others)
36. Out-of-Body experiences
    a. Overall, was this a positive or negative experience?
37. Feeling detached from the world around you
    a. Overall, was this a positive or negative experience?
38. Loss of control over behavior
    a. Overall, was this a positive or negative experience?
39. Sadness or hopelessness
40. Foggy or cloudy mind
41. Heightened degree of anger or irritability
42. Bothersome or worrisome thoughts
43. Other
    a. Overall, was this a positive or negative experience?

**Response Anchors:** None, Mild, Moderate, Extreme

***Instructions for Floatation-REST Sessions***

The following instruction set was provided to all individuals prior to their floatation-REST session for both chair-REST and pool-REST conditions: *“Thank you so much for your help in participating in this research study. Our goal is to learn more about the effects of reducing environmental stimulation on the nervous system. Throughout the day, our brain is constantly bombarded by sensory information from the external world. In this study, we aim to understand what happens when the brain gets a chance to disconnect from this constant stimulation by floating in an environment with reduced levels of light and sound, and reduced pressure on the spinal cord. While floating, try to remain still. It’s okay if you move, but just try your best not to move too much. Also, try your best not to fall asleep. We realize that you might fall asleep on occasion, but it’s important to keep in mind that our study is focused on what happens to the brain while you are awake. While most prefer to float with the lights off, the choice is yours and you are in complete control the entire time. You can float for up to 60 minutes and you are always free to stop at any time. We will turn on some music after an hour has passed. Take your time getting up, there is no rush. Before we begin, do you have any questions?”* The pool-REST preferred condition instruction set was modified to read, *“You can float for up to 120 minutes and you are always free to stop at any time. We will turn on some music after two hours have passed.”*

***Floatation-REST Environmental Preference***

For both the pool-REST and pool-REST preferred conditions, for each session participants were allowed to select from either an open or enclosed pool, and the choice frequency was calculated for each pool condition. To determine if there were differential pool preferences (open vs. enclosed pool) between the experimental conditions (pool-REST vs. pool-REST preferred), a 2x2 chi-squared test was conducted. The pool-REST condition chose to float in the open pool for 45.7% of their sessions, and the enclosed pool for the remaining 54.3% of their sessions. The pool-REST preferred participants followed a similar pattern, choosing the open pool 54.9% of the time, and choosing the enclosed pool 45.1% of the time. There were no significant differences in pool preference between the pool-REST and pool-REST preferred conditions (*X^2^* =1.85, *p* = 0.17).

**Supplementary References:**

Hembree, E. A., Foa, E. B., Dorfan, N. M., Street, G. P., Kowalski, J., & Tu, X. (2003). Do patients drop out prematurely from exposure therapy for PTSD? *J Trauma Stress* 16(6): 555-562.

Jonsson, K., & Kjellgren, A. (2016). Promising effects of treatment with flotation-REST (restricted environmental stimulation technique) as an intervention for generalized anxiety disorder (GAD): a randomized controlled pilot trial. *BMC Complement Altern Med* 16: 108.

Taylor, S., Abramowitz, J. S., & McKay, D. (2012). Non-adherence and non-response in the treatment of anxiety disorders. J Anxiety Disord 26(5): 583-589.

*Supplemental Table 1.* Event by Visit Interaction Post-hoc Comparisons.

|  | Comparison | | | | | | | | | |
| --- | --- | --- | --- | --- | --- | --- | --- | --- | --- | --- |
|  | Visit 1 vs. 2 | | Visit 1 vs. 3 | | Visit 1 vs. 4 | | Visit 1 vs. 5 | | Visit 1 vs. 6 | |
| *Event* | *p* | *d* | *p* | *d* | *p* | *d* | *p* | *d* | *p* | *d* |
| Serenity/Peacefulness | 1.00 | 0.25 | 1.00 | 0.133 | < 0.01 | 0.68 | 0.13 | 0.46 | 0.05 | 0.54 |
| Life Appreciation | 0.82 | 0.31 | 0.26 | 0.41 | 0.02 | 0.58 | 0.40 | 0.38 | < 0.01 | 0.66 |
| Refreshed | 1.00 | 0.00 | 1.00 | 0.25 | 0.05 | 0.52 | 1.00 | 0.13 | 0.02 | 0.59 |
| Relaxed | 1.00 | 0.18 | 0.29 | 0.39 | 0.05 | 0.52 | < 0.01 | 0.64 | < 0.01 | 0.66 |
| Silent Mind | < 0.01 | 0.63 | < 0.01 | 0.95 | < 0.01 | 1.08 | < 0.01 | 1.06 | < 0.01 | 0.88 |
| Pain Free Existence | 0.85 | 0.28 | < 0.01 | 0.83 | 0.43 | 0.36 | 1.00 | 0.26 | 0.48 | 0.35 |
| Feelings of Flow | < 0.01 | 0.81 | < 0.01 | 0.85 | < 0.01 | 0.61 | < 0.01 | 0.85 | < 0.01 | 0.83 |
| Dizziness | 1.00 | 0.01 | 0.46 | 0.36 | 0.84 | 0.29 | 0.04 | 0.54 | 0.25 | 0.42 |

*Note. p* indicates p-value associated with post-hoc comparison test of simple effects. *d* represents associated Cohen’s D effect size.

*Supplemental Table 2.* Event by Condition Interaction Post-hoc Comparisons.

|  | Comparison | | | | | |
| --- | --- | --- | --- | --- | --- | --- |
|  | Chair-REST vs.  Pool-REST | | Chair-REST vs.  Pool-REST Preferred | | Pool-REST vs.  Pool-REST Preferred | |
| *Event* | *p* | *d* | *p* | *d* | *p* | *d* |
| Joy/Happiness | < 0.01 | 0.69 | 0.04 | 0.34 | 0.04 | 0.35 |
| Increased Energy | < 0.01 | 0.68 | < 0.01 | 0.67 | 0.98 | 0.00 |
| Increased Focus | < 0.01 | 0.46 | < 0.01 | 0.49 | 0.85 | 0.03 |
| Serenity/Peacefulness | < 0.01 | 0.94 | < 0.01 | 1.01 | 0.63 | 0.07 |
| Empathy/Compassion | 0.01 | 0.47 | 0.42 | 0.13 | 0.04 | 0.35 |
| Life Appreciation | < 0.01 | 0.85 | < 0.01 | 0.70 | 0.29 | 0.16 |
| Refreshed | < 0.01 | 1.17 | < 0.01 | 1.25 | 0.58 | 0.08 |
| Relaxed | < 0.01 | 0.61 | < 0.01 | 1.16 | < 0.01 | 0.55 |
| Silent Mind | < 0.01 | 0.55 | < 0.01 | 0.64 | 0.53 | 0.09 |
| Pain Free Existence | < 0.01 | 0.94 | < 0.01 | 1.34 | < 0.01 | 0.40 |
| Feelings of Flow | < 0.01 | 1.24 | < 0.01 | 0.87 | 0.01 | 0.37 |
| Fear/Panic | 0 .08 | 0.33 | 0.05 | 0.38 | 0.71 | 0.05 |
| Itchiness | 0.02 | 0.42 | < 0.01 | 0.60 | 0.22 | 0.18 |
| Hallucinations | 0.31 | 0.19 | 0 .03 | 0.40 | 0.31 | 0.21 |
| Detachment | < 0.01 | 0.78 | < 0.01 | 0.81 | 0.82 | 0.03 |

*Note. p* indicates p-value associated with post-hoc comparison test of simple effects. *d* represents associated Cohen’s D effect size
